# Emergency integrative supportive care program for frail patients with advanced pancreatic cancer: A prospective GERCOR ARCAD study

**DOI:** 10.64898/2026.06.18.26355998

**Authors:** Benoit Rousseau, Marc Hilmi, Antoine Falcoz, Dewi Vernerey, Clémence Toullec, Thierry Lecomte, Aurélien Lambert, Christophe Tournigand, Véronique Guerin-Meyer, Christophe Louvet, Isabelle Trouilloud, Yves Rinaldi, Romain Coriat, Jérôme Dauba, Cindy Neuzillet, Thierry André, Jean-Baptiste Bachet, Jérôme Cros, Christelle de la Fouchardiere, Marie-Line Garcia-Larnicol, Aimery de Gramont, Pascal Hammel

**Author notes:** **Corresponding author:** Benoît Rousseau, MD, PhD, Department of Medicine - Solid Tumor Division, Mortimer B. Zuckerman Research Center, Memorial Sloan Kettering Cancer Center, 417 E 68th St, New York, NY 10065, USA.

## Abstract

**Background:** Patients with advanced pancreatic ductal adenocarcinoma (aPDAC) often experience general health decline at diagnosis due to a high-symptom burden. The optimal management of symptoms and/or poor performance status (PS) in these patients remains an unmet medical need.

**Patients and Methods:** In this multicenter study, patients with PS≥2 and pathologically confirmed or imaging-suspected aPDAC were included at first oncology visit in a personalized 14-day emergency integrative supportive care program (14-EISCP) to manage pain, nutrition, diagnostics, and stenting procedures. The primary endpoint was the 14-EISCP success in feasibility of planned procedures and clinical benefit defined as post-EISCP PS≤1, ≥5 points improvement in fatigue, pain, global health-related quality of life (HRQoL) scores (EORTC QLQ-C15-PAL), or chemotherapy initiation within 30 days.

**Results:** A total of 106 patients were included; 93 evaluable patients considered for primary endpoint analysis (median age: 76 years [68-80], PS3: 20.9%, metastases: 61.3%). The median overall survival was 4.1 months (IC95% 2.6-5.6). The 14-EISCP was successful in 59.1% (n=55) of patients, meeting the primary objective (clinically relevant). The 14-EISCP feasibility was achieved in 70.9% of cases. Post-EISCP clinical benefit was observed in 79.6% of patients, with PS improvement to 0/1 in 13.2%, HRQoL improvement in 23.9%, and chemotherapy initiation ≤30 days in 73.1%. Among evaluable patients, 17.2% received mFOLFIRINOX or gemcitabine-nab-paclitaxel, 35.4% received FOLFOX, 25.3% had gemcitabine or 5-fluorouracil alone, and 22.2% received best supportive care. In patients with PS2 at baseline, the administration of doublet/triplet chemotherapy was associated with improved overall survival compared to single-agent.

**Discussion:** These results offer a promising framework for improving outcomes in aPDAC patients, bridging the gap between symptom management and systemic therapy administration.

**Conclusions:** In patients with PS≥2 and aPDAC, the personalized 14-EISCP was feasible and lead to meaningful clinical benefit, allowing doublet or triplet chemotherapy in half the patients.

## 1. Introduction

The incidence of pancreatic ductal adenocarcinoma (PDAC) is dramatically increasing, especially in Western countries.^1^ With a 5-year overall survival (OS) rate of less than 12% and projections showing PDAC will account for 11% of cancer-related deaths by 2040, it is a major public health concern.^1,2^ Only 15%-20% of patients are eligible for a curative-intent surgical resection, and recurrence occurs in over 75% of cases after adjuvant chemotherapy.^3^ Due to the lack of early symptoms and high and prompt invasive potential of this cancer, 80% of cases are often diagnosed at an advanced stage with either metastases or locoregional extension.^4,5^

Therapeutic options for advanced PDAC (aPDAC) rely on chemotherapy and radiotherapy in patients, with only modest improvements in OS.^2,6^ The FDA has approved three regimens for metastatic PDAC: FOLFIRINOX (5-fluorouracil [5-FU], irinotecan, and oxaliplatin), NALIRIFOX (NAL-IRI 50mg/m^2^, oxaliplatin, 5-FU), and the gemcitabine plus nab-paclitaxel combination.^7–9^ However, these are primarily limited to patients with performance status (PS) ≤1 due to a less favorable toxicity profile compared to gemcitabine monotherapy. The princeps MPACT study included 65 patients (8%) with Karnofsky Index of 60 or 70 (corresponding to PS2). Subgroup analyses indicated that patients with a Karnofsky Index of 70 to 80 had a significant survival benefit with the combination regimen compared to gemcitabine alone.^8^ Real-world data suggest that 15%-30% of patients with PS2 receive chemotherapy,^10^ but their treatment options remain limited.^11^ In selected patients with PS2, gemcitabine plus nab-paclitaxel may be proposed to maximize the best chance of response.^12,13^ For patients with PS2 and/or elevated bilirubin level, only gemcitabine monotherapy can be considered. In patients with PS≥3 and limited life expectancy, chemotherapy may be inappropriate, and best supportive care (BSC) should be considered the most relevant option. PDAC frequently causes severe symptoms such as fatigue, pain, and malnutrition. Many patients with aPDAC are referred to specialized gastrointestinal cancer centers late (i.e. several weeks after first medical contact).^14^ These factors impair prognosis and may preclude administration of chemotherapy. In contrast, improved health-related quality of life (HRQoL) correlates with better survival,^15–17^ yet patients with frailty are often excluded from clinical trials. Early palliative care may positively impact prognosis and symptom management.^18,19^ Therefore, treatment decisions in this population must balance symptom control with overall prognosis. Optimal care for patients with symptomatic aPDAC and/or poor PS remains an unmet medical need.

This prospective multicenter study evaluated the feasibility and clinical impact of implementing an emergency integrative supportive care program (EISCP), including pain and nutritional management, diagnostic biopsies, and therapeutic endoscopy, within 14 days, with the goal of improving patients’ condition, health-related quality of life (HRQoL), and timely access to optimal chemotherapy. Additionally, a preplanned secondary endpoint of the study was to evaluate the impact of the 14-day EISCP (14-EISCP) success and chemotherapy type on patient prognosis.

## 2. Methods

### 2.1 Trial design and participants

This is a multicenter, prospective study that enrolled patients with a PS≥2 and pathologically confirmed or highly suspected aPDAC based on imaging, with clinico-biological features precluding initial treatment with FOLFIRINOX or gemcitabine-nab-paclitaxel. Patients were included by the local investigator at their first oncology visit (V1) if standardized clinical assessments indicated a need for at least two predefined components of the 14-EISCP. These components were: (i) a specialized supportive care consultation for pain guided by Visual Analogue Scale (VAS) assessment, and/or nutrition per ESPEN guidelines using validated items (recent weight loss, BMI, serum albumin, oral intake)^20^; (ii) pathological confirmation if no tissue diagnosis was available (endoscopy-guided or percutaneous biopsies, ascites cytology); and (iii) endoscopic stenting of the bile duct/duodenum when obstruction was confirmed. The investigator selected procedures and emailed them to the sponsor for validation, which set the program start date and 14-day timeframe. The 14-EISCP began as soon as aPDAC was suspected at first consultation. Patients with a prior cancer were excluded.

At the end of the 14-EISCP, a second visit (V2) documented supportive care, completed procedures, reassessed PS, administered the EORTC QLQ-C15-PAL, and determined therapeutic allocation. Patients with PS≤1, eligible for FOLFIRINOX or gemcitabine-nab-paclitaxel, entered Group-1. Patients with persisting PS≥2 or clinico-biological features precluding these regimens (bilirubin >1.5× ULN, organ dysfunction, or comorbidities contraindicating intensive chemotherapy) entered Group-2. Group-1 received mFOLFIRINOX or gemcitabine-nab-paclitaxel, Group-2 received investigator’s choice among mFOLFOX6, single-agent chemotherapy (gemcitabine/fluoropyrimidine), or BSC. Chemotherapy was allowed without pathological confirmation, if at least one attempt to obtain tissue failed. Eligibility and group allocation were determined by a multidisciplinary tumor board, with any uncertainty resolved with the coordinating investigator. One-month post-V1, a third visit (M1) was conducted, followed by bimonthly visits. These visits included a clinical exam with assessment of PS and QLQ-C15-PAL. All participants provided written informed consent in accordance with International Council for Harmonization Good Clinical Practice (ICH/GCP), and national or local regulations before registration. This study followed the CONSORT reporting guidelines. This study was approved by the Ethic review board (RCB 2016-A01077-44, NCT02979483).

### 2.2 Patient assessment and outcomes

The primary endpoint was the 14-EISCP success rate, defined by a co-primary criteria including feasibility of the 14-EISCP and the clinical benefit at the end of the program. The 14-EISCP was considered feasible if all planned procedures were completed within 14 days (±2 days) post-V1. If a planned procedure was incomplete or delayed beyond 14 ±2 days, the feasibility was deemed as failed.

Clinical benefit of the 14-EISCP was defined as: (i) improvement of PS from ≥2 to 1 or 0, (ii) a ≥5 points improvement in either fatigue, pain, or global health, as evaluated by QLQ-C15-PAL^21^ without worsening of PS and other QLQ-C15-PAL dimensions (fatigue, pain, and global heath), or (iii) initiation of chemotherapy within 30 days after V1. Failure to meet any of these criteria resulted in clinical benefit being classified as unsuccessful. Secondary endpoints included OS from V1 and V2, the delay between V1 and chemotherapy administration, and change in HRQoL from V1 to V2 and to M1 (based on QLQ-C15-PAL dimensions). HRQoL was assessed at V1, V2, and M1 using EORTC QLQ-C15-PAL.

The 14-EISCP was considered successful only if both feasibility and clinical benefit criteria were met.

### 2.3 Statistical analysis

All analyses were performed using SAS version 9.4 (SAS Institute, Cary, North Carolina, USA) and R software version 4.1.1 (R Development Core Team, Vienna, Austria).

Considerations of determining the appropriate sample size were based on the success rate of the 14-EISCP, with 50% considered uninteresting and 64% as satisfactory. Based on these hypotheses, an A’Hern one-step design^22^ was used with unilateral alpha type I error of 5%. Due to the COVID-19 pandemic and the resulting low recruitment rate, the protocol was amended in May 2021. As a result, the statistical power decreased from 90% to 85% to reduce the number of patients needed to meet the primary objective, from 105 to 93 evaluable patients. In this amended protocol, 98 patients needed to be included, with at least 55 (59%) patients meeting the success rate definition. The primary endpoint analysis population was estimated to consist of the first 93 evaluable patients.

The primary objective was assessed in the modified intention-to-treat (mITT) population, corresponding to metastatic and locally advanced patients assessable for the 14-EISCP success, regardless of inclusion/exclusion criteria.

Patient and tumor characteristics were described for the overall population, with categorical variable expressed as percentages and continuous variables reported as mean with standard deviations. OS and progression-free survival (PFS) were estimated using the Kaplan-Meier method and presented as medians with 95% CI). All survival analyses were done on a population of all included patients with survival data available and confirmation of aPDAC. Hazard ratios (HRs) were calculated using Cox proportional hazard regression models. The association of baseline parameters with outcomes was first examined using an univariable Cox analysis, followed by inclusion of variables with a *P* <.15 in a multivariable Cox regression model. Follow-up was estimated using the reverse Kaplan-Meier method and was reported as the median with its 95% CI. For the analysis evaluating the association between chemotherapy initiation within 30 days and OS, we used a landmark approach with survival measured from day 15 (after completion of the 14-day intervention period) to avoid a time bias. The student’s t-test compared continuous variables, while Chi-square tested categorial variables. For paired proportions, Wilcoxon signed-rank test was used. HRQoL was described at V1, V2, and M1 for the targeted dimensions (global health, fatigue, pain), and change from baseline was tested using the Wilcoxon signed-rank test.

For the primary objective, a one-sided type I error of 5% and a statistical power of 85% were planned. All other *P*-values were two-sided and considered exploratory, without any correction for multiple testing.

## 3. Results

### 3.1 Patient characteristics

Out of the 106 patients included in the study, seven were excluded from evaluation due to insufficient feasibility data (n=3), clinical benefit data (n=3), and study discontinuation based on patient preference (n=1; Supplemental eFigure 1). Patient characteristics are presented in Supplemental eTable 1. In the primary endpoint population (n=93), 77.4% had a PS2 (n=72) and 22.6% had a PS of 3 (n=21), and 61.3% of patients had metastases (n=57).

The median age was 76.0 years (interquartile range 68.0-79.9), and the median delay between the first symptoms to V1 was 55.0 days (25.0-94.0). Planned interventions at baseline (V1) appear in Supplemental eTable 1. Most patients required specialized consultation in the institutional palliative care unit for nutrition management (87.1%, n=81) and pain management (67.7%, n=63). Pathological diagnosis was performed before V1 in 48.4% of cases (n=45) and 51.6% of patients required pathological confirmation after V1 (n=48). Two patients died before receiving the pathological diagnosis by biopsy. Among the 46 patients who underwent pathological diagnosis after inclusion, 91.3% (n=42) had pathological confirmation of aPDAC. One patient had an unsuccessful pathological assessment, and three patients had another final diagnosis than PDAC (two benign tumors of the pancreas and one sarcoma).

Stent placement in the biliary and duodenal tracts was required in 16.1% (n=15) and 8.6% (n=8) of cases, respectively.

### 3.2 Primary endpoint

The 14-EISCP was a success (i.e. feasible with a clinical benefit) in 59.1% (90% CI 0.50-0.68) of patients (n=55), meeting the primary objective (Table 1). Considering each component defining success, the 14-EISCP feasibility was achieved in 70.9% of patients (n=66), and clinical benefit was observed in 79.6% (n=74; Table 1 and Supplemental eFigure 1).

**Table 1.**
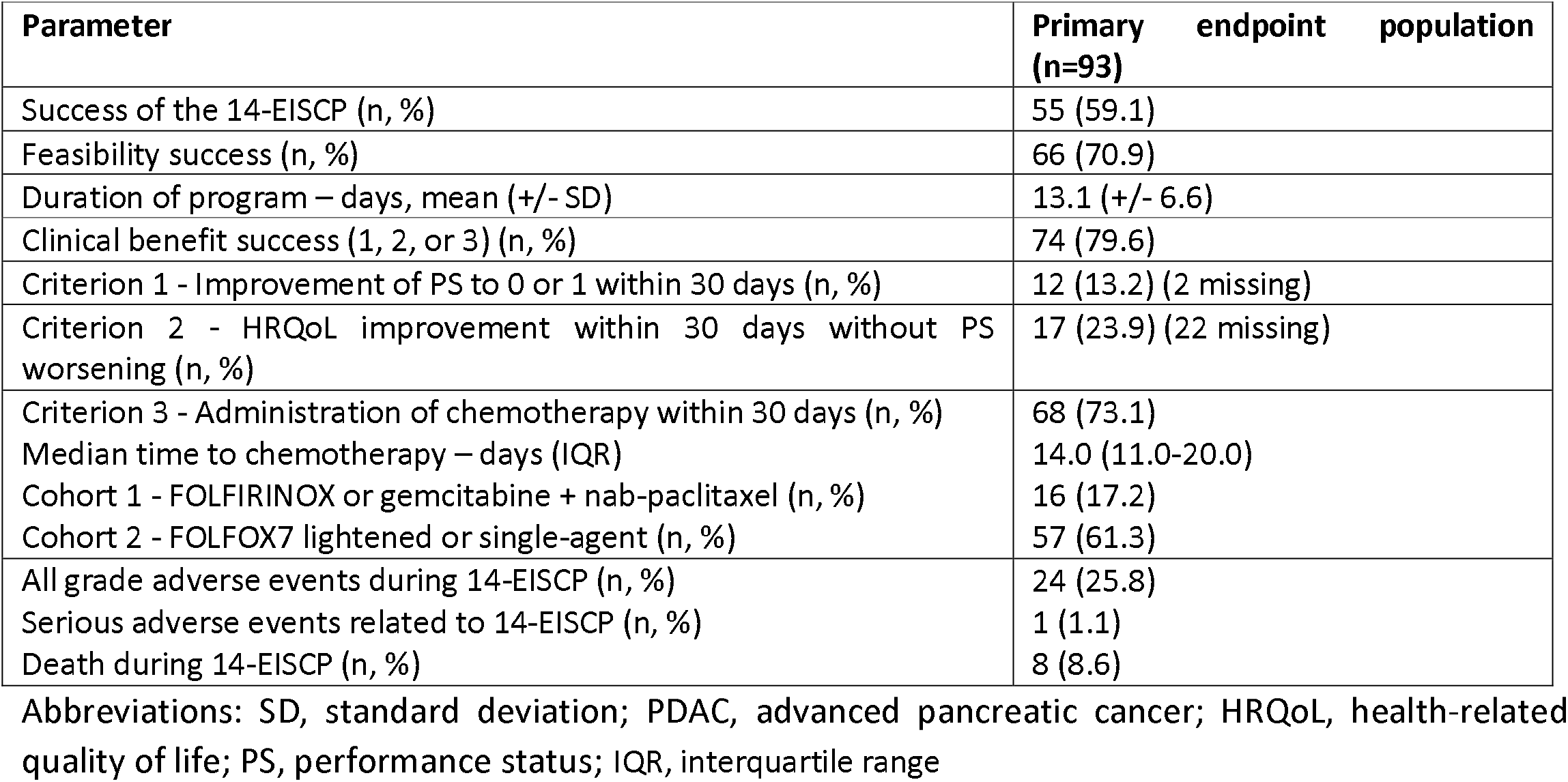
Results of the personalized 14-day emergency integrative supportive care program (14-EISCP)

| Parameter | Primary endpoint population (n=93) |
| --- | --- |
| Success of the 14-EISCP (n, %) | 55 (59.1) |
| Feasibility success (n, %) | 66 (70.9) |
| Duration of program – days, mean (+/- SD) | 13.1 (+/- 6.6) |
| Clinical benefit success (1, 2, or 3) (n, %) | 74 (79.6) |
| Criterion 1 - Improvement of PS to 0 or 1 within 30 days (n, %) | 12 (13.2) (2 missing) |
| Criterion 2 - HRQoL improvement within 30 days without PS worsening (n, %) | 17 (23.9) (22 missing) |
| Criterion 3 - Administration of chemotherapy within 30 days (n, %) | 68 (73.1) |
| Median time to chemotherapy – days (IQR) | 14.0 (11.0-20.0) |
| Cohort 1 - FOLFIRINOX or gemcitabine + nab-paclitaxel (n, %) | 16 (17.2) |
| Cohort 2 - FOLFOX7 lightened or single-agent (n, %) | 57 (61.3) |
| All grade adverse events during 14-EISCP (n, %) | 24 (25.8) |
| Serious adverse events related to 14-EISCP (n, %) | 1 (1.1) |
| Death during 14-EISCP (n, %) | 8 (8.6) |

Abbreviations: SD, standard deviation; PDAC, advanced pancreatic cancer; HRQoL, health-related quality of life; PS, performance status; IQR, interquartile range

Regarding feasibility, the completion rates of planned procedures within the 14-EISCP were 88% for nutritional care, 81% for pain management, 76% for anatomopathological diagnosis, 60% for biliary stenting, and 50% for duodenal stenting (Supplemental eFigure 2A). Failures in the 14-EISCP feasibility were mainly due to one procedure (51.9%, n=14/27) or two unperformed procedures (37.0%, n=10/27, Supplemental eTable 2).

During the 14-EISCP, eight patients died (8.6%, n=8/93). Cancer caused death in five cases. Of the remaining three, two were non-procedures-related (one due to sepsis related to biliary stenting before inclusion and one due to hyperglycemia linked to parenteral nutrition before inclusion), and one was procedure-related (a sudden death, likely from pulmonary embolism after discontinuing anticoagulant for a liver biopsy) (Table 1).

Considering the three criteria defining the clinical benefit (Table 1), 13.2% (n=12/91) of patients had an improvement in PS to 0 or 1, 23.9% (n=17/71) of patients showed an improvement of their HRQoL without worsening of their PS, and 73.1% (n=68/93) of patients received chemotherapy within 30 days. Clinical benefit did not differ according to feasibility (78.1% for feasible group vs 66.7% fornon-feasible group, *P*=.64). At the end of the 14-EISCP, 63.2% (n=55) of patients remained at PS2, 16.1% (n=14) were at PS3, and 4.6% at PS4 (n=4). Among patients with available HRQoL questionnaires, pain levels decreased significantly at both V2 and M1, (*P*=.006 and *P*=.002, respectively). Fatigue also decreased at V2 (*P*=.02) and stabilized at M1 (*P*=.50) compared to baseline (Supplemental eFigure 3).

In the evaluable cohort for the primary endpoint, 80.6% (n=75/93) of patients received chemotherapy after the 14-EISCP with 73.1% (n=68/93) within 30 days. Chemotherapy was administered within 30 days in 53.8% (n=50/93) of patients who did not experienced improvements in PS or HRQoL, thus meeting the predefined criteria for clinical benefit (Supplemental eFigure 2B).

At the end of the 14-EISCP, patients’ therapeutic allocationwas the following: 17.2% entered Group-1 (n=16/93) receiving FOLFIRINOX or gemcitabine-nab-paclitaxel, while 61.3% (n=57/93) entered Group-2 receiving mFOLFOX6 or single-agent chemotherapy (gemcitabine or 5-FU). Two patients in Group-2 subsequently received mFOLFIRINOX.

### 3.3 Survival outcomes

#### a) Primary endpoint analysis

Among the 96 evaluable patients for survival analyses, median OS (mOS) from V1 was 4.1 months (CI 95% 2.8-5.7). Regarding the co-primary endpoints, neither the success nor feasibility of the 14-EISCP was associated with survival (*P*=.21 and *P*=.38, respectively; Figure 1A-B). However, mOS was numerically longer in patients with clinical benefit (4.6 months, 95% CI 3.3-6.6) compared to those without clinical benefit (2.7 months, 95% CI 1.8-6.1), though this difference did not reach statistical significance (P=.06; Figure 1C).. In the landmark analysis from day 15 (V2), early chemotherapy initiation (within 30 days from inclusion at V1) was significantly associated (*P*=.01) with improved survival compared to later start or BSC (Figure 1D). Patients who died during the intervention period were excluded from this survival analysis (N=8).

**Figure 1.**
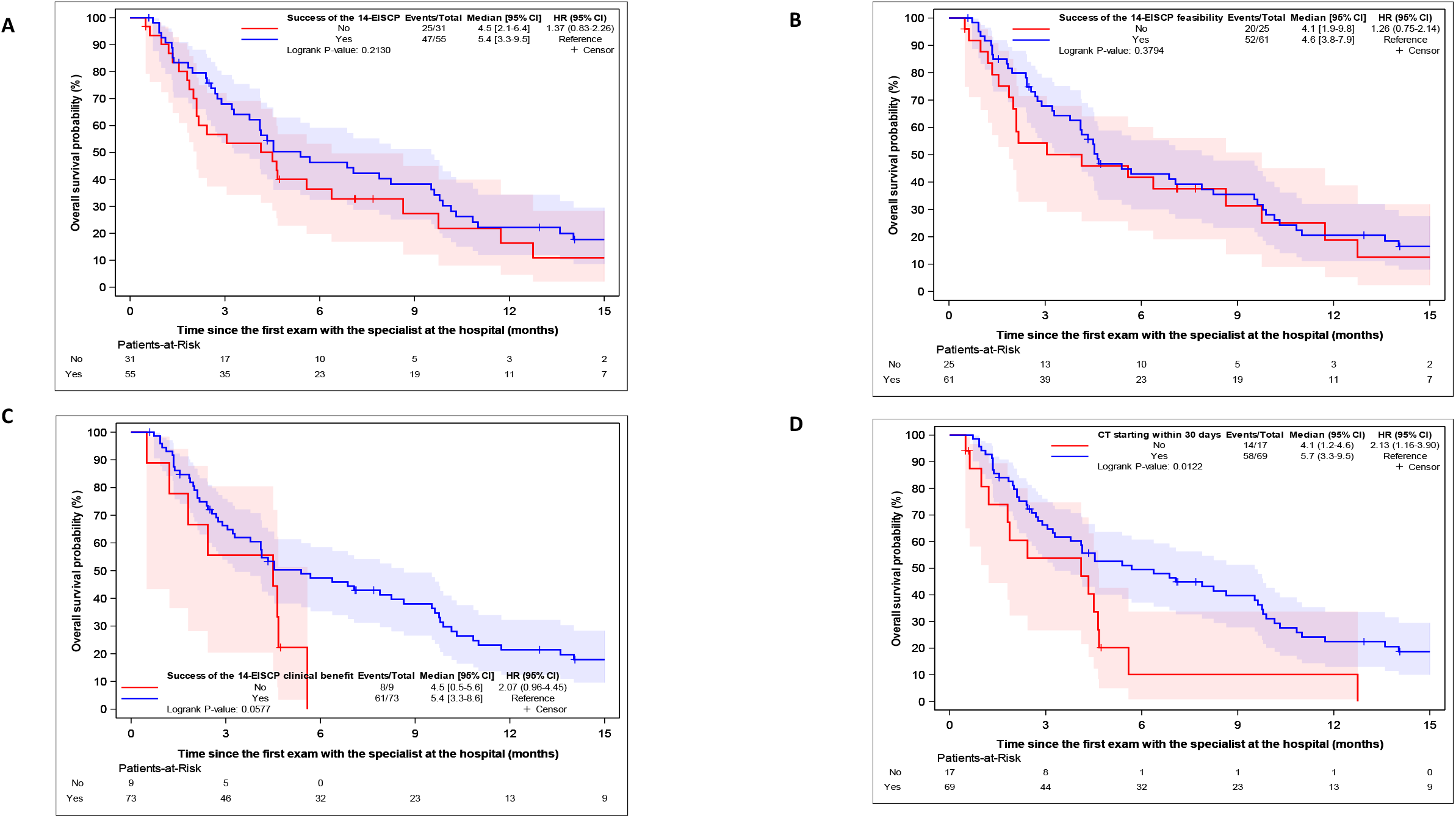
Overall survival in evaluable patients according to the primary endpoint. Overall survival according to success (A), feasibility (B), clinical benefit of the integrative supportive care program (C) and chemotherapy administration within 30 days (D) (landmark analysis from 15 days post inclusion).

#### b) Exploratory analysis

When comparing OS by PS at V1, patients with PS2 had a mOS of 4.5 months, whereas those with PS3 had a markedly shorter mOS of 2.1 months. Among patients with PS2 at V1, mOS was 7.9 months for those who received chemotherapy (n=56) vs. 4.3 months for those managed with BSC (n=15) (*P*=.03; Figure 2A). When comparing survival of PS2 patients according to chemotherapy regimen, OS was not statistically different between those receiving FOLFIRINOX/gemcitabine-nab-paclitaxel and those receiving mFOLFOX6 (Figure 2B). However, patients receiving single-agent chemotherapy had inferior OS compared with those receiving mFOLFOX6/FOLFIRINOX/gemcitabine-nab-paclitaxel (*P*=.02, Figure 2C). In contrast, among patients with poor PS 3-4 at V1, receipt of chemotherapy was not associated with an improvement in overall survival (P=.20, Figure 2D).

**Figure 2.**
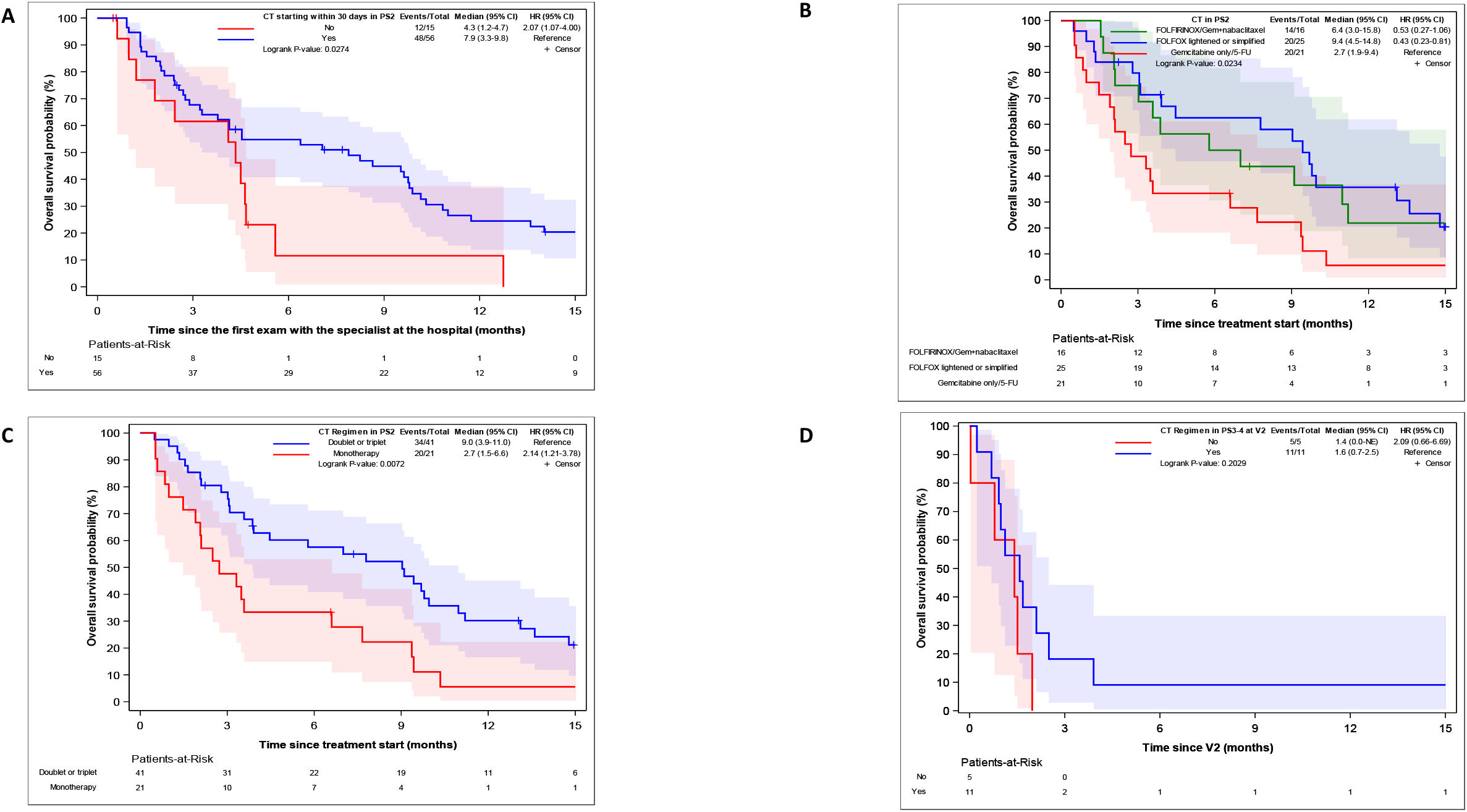
Survival in evaluable patients according to PS and treatment. (A) Overall survival in patients with PS2 according to receipt of chemotherapy vs. best supportive care. (B) Overall survival in patients with PS2 according to chemotherapy regimen: FOLFIRINOX or gemcitabine–nab-paclitaxel vs. FOLFOX vs. single-agent chemotherapy (gemcitabine or 5-FU). (C) Overall survival in patients with PS2 according to chemotherapy regimen: doublet or triplet vs. single-agent chemotherapy. (D) Overall survival in patients with PS3 according to receipt of chemotherapy vs. best supportive care.

In the multivariate model (Table 2), significative independent prognostic factors for OS were age >75 years (HR=0.51, 95% CI, 0.28-0.93]; *P*=.03), metastatic stage (HR=1.79, 95% CI, 1.05-3.04, *P*=.03), decreased global quality of life at V2 <66.7 (HR=1.76, 95% CI, 1.01-3.08, *P*=.05), and chemotherapy regimen (HR=0.29, 95% CI, 0.11-0.76; *P*=.01 for mFOLFOX6, HR=0.15, 95% CI, 0.04-0.55; *P*<.01 for FOLFIRINOX/gemcitabine-nab-paclitaxel). Survival was numerically longer for patients receiving single-agent chemotherapy compared with best supportive care (HR=0.39, 95% CI 0.15-1.03, P=.06), though this difference did not achieve statistical significance. When comparing QoL trajectories, patients who initiated chemotherapy showed significant improvements from V2 in several domains compared with those who did not receive chemotherapy (Supplementary Table 4). Specifically, emotional functioning improved, and symptom burden decreased for pain, dyspnea, and insomnia, whereas no comparable improvement was observed in the no-chemotherapy or delayed-chemotherapy group.Baseline characteristics were compared between patients who did and did not receive chemotherapy within 30 days (Table 3). No significant differences were observed in prognostic variables between the two groups. However, patients who received chemotherapy within 30 days were less likely to have a biliary stent compared with those who did not (0.7% vs 26.9%, p = 0.015), suggesting that the timing of biliary stent placement may influence the initiation of chemotherapy without reflecting differences in baseline patient characteristics.

**Table 2.** Univariate and multivariate analyses of overall survival from V2.

|  | Univariable analysis |  |  |  |  | Multivariable analysis |  |  |
| --- | --- | --- | --- | --- | --- | --- | --- | --- |
|  | N | events | HR | 95% CI | P-value | HR | 95% CI | P-value |
| **N (Event) – C-index** |  |  |  |  |  | **80 (67) – 0.7163** |  |  |
| **Age at inclusion (years)** |  |  |  |  |  |  |  |  |
| ≤75 | 36 | 31 | 1 |  |  | 1 |  |  |
| >75 | 50 | 42 | 0.682 | 0.425-1.094 | 0.1126 | 0.510 | 0.279-0.933 | 0.0287 |
| **Disease stage** |  |  |  |  |  |  |  |  |
| Locally advanced | 33 | 25 | 1 |  |  | 1 |  |  |
| Metastatic | 53 | 48 | 1.575 | 0.968-2.561 | 0.0673 | 1.789 | 1.054-3.035 | 0.0310 |
| **Chemotherapy regimen** |  |  |  |  |  |  |  |  |
| No chemotherapy | 10 | 8 | 1 |  |  | 1 |  | 0.0309 |
| Gemcitabine only/5-FU | 24 | 22 | 0.393 | 0.171-0.903 | 0.0277 | 0.386 | 0.145-1.031 | 0.0577 |
| FOLFOX | 35 | 29 | 0.234 | 0.102-0.536 | 0.0006 | 0.285 | 0.107-0.756 | 0.0117 |
| FOLFIRINOX/Gemcitabine+nab-paclitaxel | 17 | 14 | 0.214 | 0.086-0.529 | 0.0008 | 0.145 | 0.038-0.551 | 0.0046 |
| **ECOG PS at visit 2 (V2)** |  |  |  |  |  |  |  |  |
| 1 | 13 | 11 | 1 |  | 0.0005 | 1 |  | 0.0463 |
| 2 | 55 | 44 | 1.150 | 0.591-2.238 | 0.6815 | 0.734 | 0.322-1.675 | 0.4628 |
| 3-4 | 16 | 16 | 3.550 | 1.610-7.830 | 0.0017 | 1.662 | 0.581-4.754 | 0.3435 |
| **Global quality of life at visit 2 (V2)** |  |  |  |  |  |  |  |  |
| ≥66.7 | 31 | 23 | 1 |  |  | 1 |  |  |
| <66.7 | 50 | 45 | 2.118 | 1.263-3.552 | 0.0045 | 1.762 | 1.009-3.078 | 0.0465 |

**Table 3.**
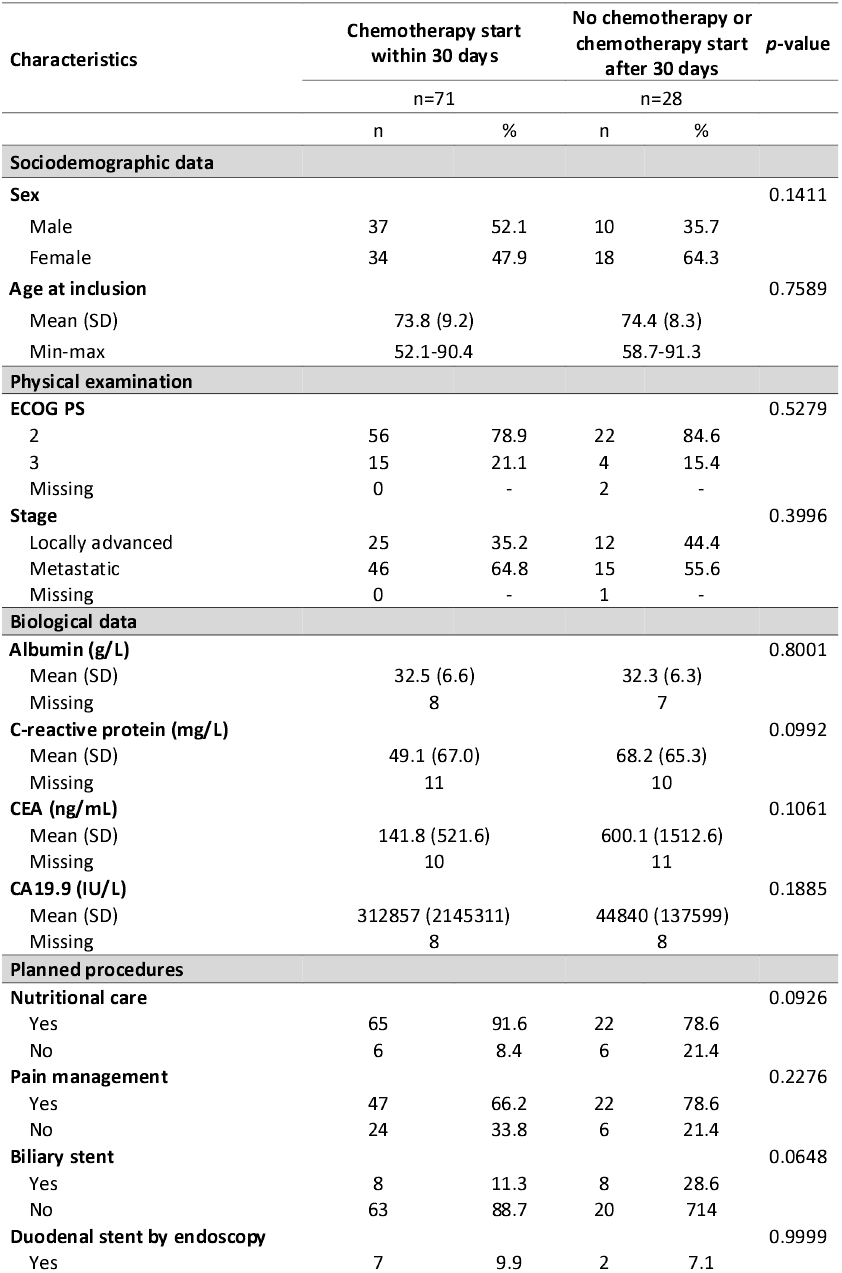
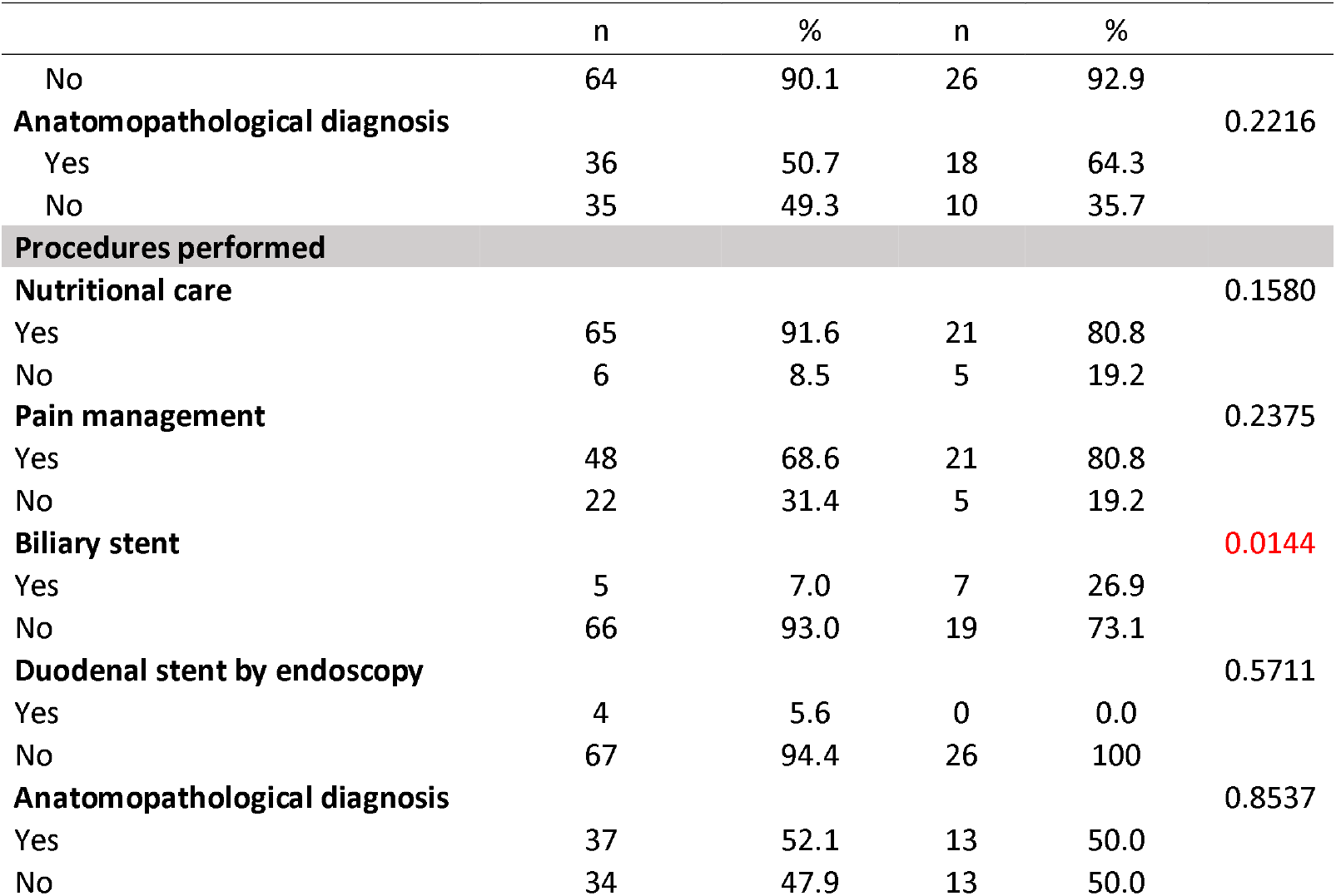
Patients’ characteristics at V1 according to chemotherapy start in evaluable population for the primary endpoint analysis (n=99).

## 4. Discussion

This multicenter, prospective study suggests that implementing a personalized 14-EISCP for patients with aPDAC and poor PS≥2 is both feasible and clinically beneficial. This innovative program was defined using composite criteria, including the feasibility of systemic chemotherapy and potential improvements in PS and HRQoL within 30 days. Feasibility was achieved in 70.9% of patients, and 59.1% met the program’s success criteria. Although feasibility was not directly associated with improved OS, chemotherapy initiation within 30 days was significantly correlated with better survival outcomes. Patients who received only BSC instead of chemotherapy within 30 days of inclusion experienced worsened PS or infections, underlining the urgency of treating symptomatic aPDAC as an oncologic emergency. Eighteen patients did not improve sufficiently to receive chemotherapy, with eight dying within 14 days, highlighting the aggressive nature of aPDAC and the need for timely referral and intervention. Similarly, a single-center retrospective cohort study of 477 PDAC patients showed a median delay of 2 months between symptom onset and diagnosis.^14^ This finding reflects the aggressiveness of this disease, reinforcing the need for rapid treatment initiation.

The mOS was 7.1 months with FOLFIRINOX or gemcitabine-nab-paclitaxel, aligns with previous trials showing similar outcomes in patients with PS2^12^. In comparison, clinical trials in PS0/1 patients treated with FOLFIRINOX and gemcitabine-nab-paclitaxel had median OS of 11.1/8.5 months, respectively.^8,23^ These findings suggest that frail patients with aPDAC and PS≥2 at diagnosis who show improvement after supportive care might benefit from doublet or triplet chemotherapy, similar to those with better initial PS. In this study, monotherapy with 5-FU or gemcitabine appeared less effective than doublet chemotherapy and showed only a trend for better survival compared to BSC. In the EA2186 study, despite utilizing attenuated chemotherapy regimens (gemcitabine/nab-paclitaxel vs. 5-FU/leucovorin/liposomal irinotecan), no significant survival benefit was observed in older, vulnerable adults with newly diagnosed aPDAC.^24^

Although early initiation of chemotherapy was significantly associated with improved OS, this finding must be interpreted cautiously. In non-randomized studies, unmeasured confounders such as subtle differences in baseline frailty, comorbidities, or center-level practices may have influenced decision and timing of treatment. While chemotherapy initiation in France is guided by clinical criteria rather than patient preference, variability in physician judgment may still contribute to selection bias. Additionally, patients who deteriorated rapidly may have been excluded from chemotherapy due to early death or clinical decline, introducing immortal time bias. These limitations underscore the need for randomized controlled trials to validate the causal relationship between supportive interventions, early chemotherapy access, and survival outcomes in frail patients with aPDAC. Nonetheless, our results align with other data showing that early diagnosis and short time-to-treatment are associated with better survival in aPDAC.^25^

While chemotherapy initiation was one of the predefined criteria for clinical benefit, we acknowledge that its inclusion may be controversial. In frail patients with poor PS, initiating systemic treatment may not be linked to any clinically relevant benefit, especially given the short median OS observed. However, noticeably in our study population, all patients were initially considered unfit for intensive chemotherapy. Therefore, the ability to receive chemotherapy within 30 days, after structured supportive care intervention, was interpreted as a surrogate for clinical improvement and functional stabilization.

Importantly, clinical benefit also included objective functional and symptomatic improvements (PS improvement or ≥5-point HRQoL gain without deterioration), providing a broader, clinically meaningful definition. By day 30, improvements in HRQoL occurred in 23.9% of patients, with significant reductions in fatigue and pain, underscoring the program’s effectiveness in alleviating symptom burden. The intervention created a window of opportunity for systemic chemotherapy by addressing debilitating symptoms early. Furthermore, longitudinal QoL analyses demonstrated that patients who initiated chemotherapy experienced improvements in emotional functioning and reductions in pain, dyspnea, and insomnia, whereas no comparable improvement was observed in patients who did not receive chemotherapy. These findings suggest that, in selected patients, systemic treatment may contribute to symptom control rather than exacerbate symptom burden. To further address this topic, we performed PS-stratified survival analyses. Among patients with PS2 at baseline, chemotherapy initiation was associated with a significant improvement in OS, supporting its clinical relevance in this subgroup. In contrast, among patients with PS 3–4, chemotherapy initiation was not associated with improved survival (Figure 2D), indicating that in the frailest patients, functional and symptomatic outcomes may be more clinically meaningful than access to systemic therapy.

Our observational study design does not permit establishment of causality between the supportive care intervention and survival outcomes. While we observed a significant association between early chemotherapy initiation and improved survival, several factors limit causal inference. First, selection bias is inherent: patients who responded favorably to supportive care may have had more favorable tumor biology or better physiological reserves independent of the intervention. Second, immortal time bias cannot be fully eliminated despite our landmark analysis; patients surviving to receive chemotherapy are inherently selected. Third, confounding by indication affects treatment decisions based on clinical trajectory and subjective assessments difficult to quantify. Finally, without a randomized control group, we cannot isolate the intervention’s contribution from spontaneous improvement or disease heterogeneity. Therefore, while our findings are hypothesis-generating and suggest potential benefit from structured supportive care, definitive demonstration of causal survival benefit would require a randomized trial, ethically and practically challenging given the rapid deterioration in this population (median OS 4.1 months).

Despite overall success, the program faced challenges. The feasibility of implementing all planned procedures was limited (29.1%), mainly due to logistical issues or rapid disease progression. Future efforts should focus on earlier referrals, stronger coordination, and digital tools for real-time HRQoL monitoring. While eligibility and allocation decisions were made by investigators, they were systematically reviewed in multidisciplinary tumor boards using predefined criteria. This approach mirrors real-world care pathways and supports the pragmatic design of the study. Nevertheless, our observational design precludes establishing causality between the intervention and survival outcomes. The observed survival association may reflect patient selection, unmeasured confounding, or guarantee-time bias rather than true intervention effect. Independent central adjudication could further minimize such bias and would be appropriate for a future randomized trial.

In conclusion, a personalized 14-EISCP is feasible for patients with newly diagnosed aPDAC and ECOG PS ≥2, enabling timely chemotherapy access in 73.1% of patients. While early chemotherapy initiation was associated with improved survival, our observational design does not establish causality. These findings support the value of structured supportive care pathways and provide hypothesis-generating data warranting validation in controlled studies.

## Data Availability

All data produced in the present study are available upon reasonable request to the authors

## Funding and acknowledgements

This work is supported by a National Cancer Institute/ National Institutes of Health Cancer Center Support grant to Memorial Sloan Kettering Cancer Center (P30 CA008748), MSKCC T32-CA009512,, DOD CDA CA230829/HT94252410897 (BR), The Society of Immunotherapy of Cancer (SITC) Dr Steve Rosemberg Scholarship (BR). This work was funded by the ARCAD Foundation.

## Authors’ contributions

Conceptualization: BR, PH

Methodology: BR, AF, DV, CL, MGL, AG, PH

Data curation: BR, CT, TL, AL, CT, VGM, CL, IT, YR, JC, RC, JD, CN, TA, JBB, FDLF, CF

Formal analysis: BR, MH, AF, DV, AG, PH

Investigation: BR, CT, TL, AL, CT, VGM, CL, IT, YR, RC, JC, JD, CN, TA, JBB, FDLF, CF

Validation: all

Writing-original draft: BR, MH

Writing-review and editing: all

