## Supplementary data for "Emergency integrative supportive care program for frail patients with advanced pancreatic cancer: A prospective GERCOR ARCAD study"

**Supplementary eTable 1. Patient characteristics at inclusion.**

| **Characteristics** | **Global population (n=106)** | **Evaluable population (n=99)** | **Primary endpoint population (n=93)** |
| --- | --- | --- | --- |
| Sex – Female, % (n) | 52.8 (n=56) | 52.5 (n=52) | 53.8 (n=50) |
| Age – Years, mean +/- SD | 74.3 +/- 8.9 | 74.0 +/- 8.9 | 73.8 +/- 8.9 |
| ECOS PS % (n)  2  3 |  |  |  |
|  | 79.8 (n=83) | 80.4 (n=78) | 79.1 (n=72) |
|  | 20.2 (n=21) | 19.6 (n=19) | 20.9 (n=19) |
| Stage, % (n)  Locally advanced  Metastatic |  |  |  |
|  | 31.1 (n=32) | 37.8 (n=37) | 38.7 (n=36) |
|  | 63.8 (n=67) | 62.2 (n=61) | 61.3 (n=57) |
| Albumin – g/L, mean +/- SD | 32.2 +/- 6.3 | 32.5 +/- 6.5 | 32.4 +/- 6.5 |
| C-reactive protein – mg/L, mean +/- SD | 55.4 +/- 65.5 | 53.5 +/- 66.7 | 54.6 +/- 68.4 |
| LDH IU/L, mean +/- SD | 287.8 +/- 161.0 | 280.8 +/- 151.1 | 278.9 +/- 148.6 |
| CA19-9 IU/L, median, IQR | 2,253 (160-18,000) | 1,656 (148-14523) | 1,490 (160-14,523) |
| Anorexia (%, n) | 74.0 (n=77) | 76.3 (n=74) | 76.9 (n=70) |
| Delay between first symptoms and V1, mean +/- SD |  |  | 55.0 (25.0-94.0) |
| Planned procedures, % (n)  Pathological diagnosis  Nutritional care  Pain management  Biliary stent  Duodenal stent |  |  |  |
|  | 48.1 (n=51) | 50.5 (n=50) | 51.6 (n=46) |
|  | 88.7 (n=94) | 87.9 (n=89) | 87.1 (n=81) |
|  | 68.9 (n=73) | 69.7 (n=69) | 67.7 (n=63) |
|  | 15.1 (n=16) | 16.2 (n=16) | 16.1 (n=15) |
|  | 8.5 (n=9) | 9.1 (n=9) | 8.6 (n=8) |

**Supplementary eTable 2. Failed procedures within the 14 days EISCP relative to planned procedures in the primary endpoint population (n=93).**

| **Number of procedures failed** | n | % |
| --- | --- | --- |
| 1 | 14 | 15.1 |
| Nutritional care | 3 | 3.2 |
| Pain management | 3 | 3.2 |
| Biliary stent | 2 | 2.2 |
| Duodenal stent | 1 | 1.1 |
| Anatomopathological diagnosis by biopsy | 5 | 5.4 |
| 2 | 10 | 10.8 |
| Nutritional care – Pain management | 4 | 4.3 |
| Nutritional care – Anatomopathological diagnosis by biopsy | 1 | 1.1 |
| Pain management - Anatomopathological diagnosis by biopsy | 2 | 2.2 |
| Biliary stent – Duodenal stent | 2 | 2.2 |
| Biliary stent – Anatomopathological diagnosis by biopsy | 1 | 1.1 |
| 3 | 2 | 2.2 |
| Nutritional care – Pain management – Anatomopathological diagnosis by biopsy | 1 | 1.1 |
| Pain management - Biliary stent – Duodenal stent | 1 | 1.1 |
| 4 | 0 | 0.0 |
| 5 | 1 | 1.1 |

**Supplementary Figure e1. Feasibility and clinical benefit of the 14-EISCP.**

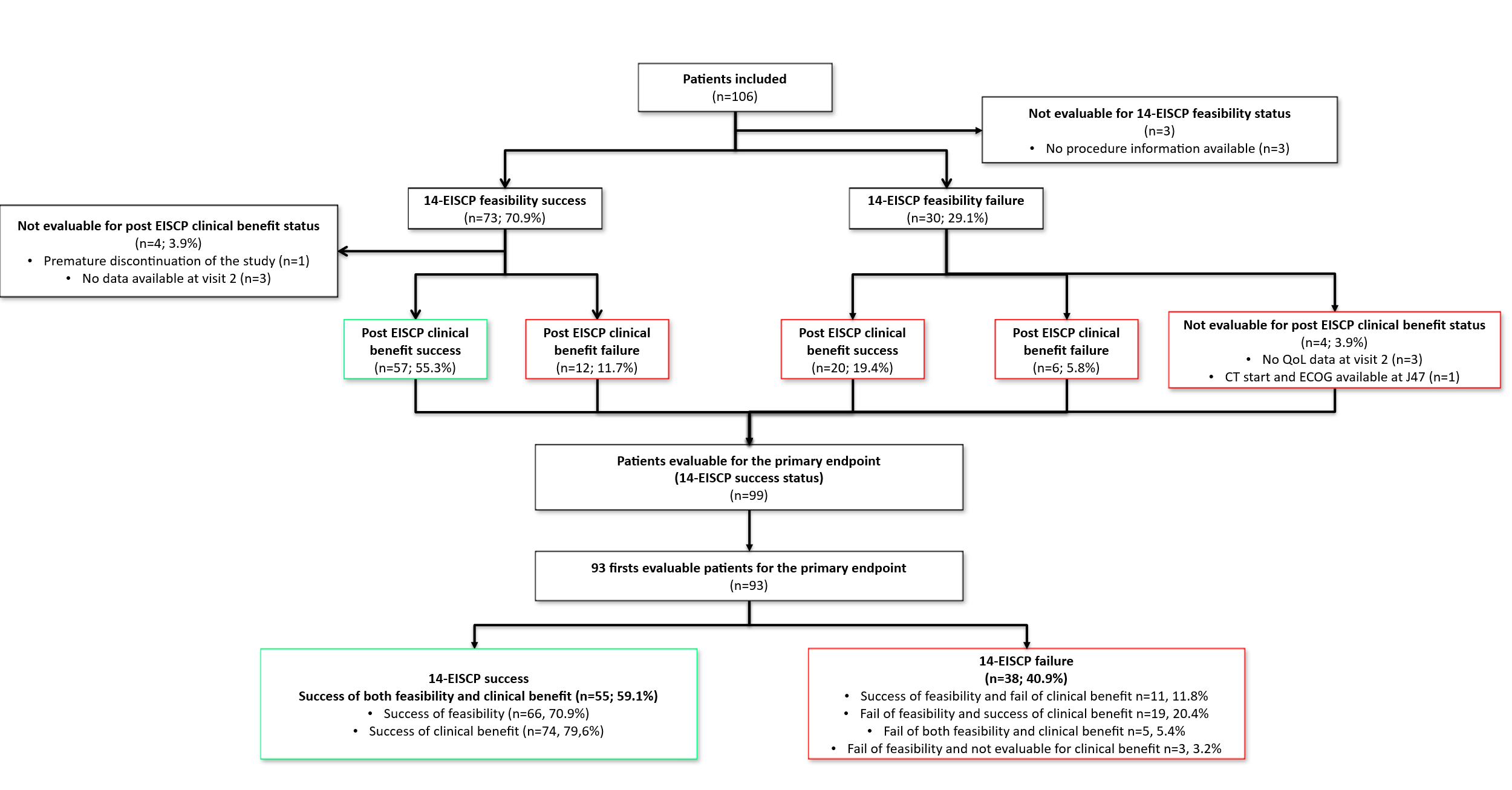

**Supplementary eFigure 2. Completion of planned procedures (A) and overlap among criteria defining the clinical benefit (B).**

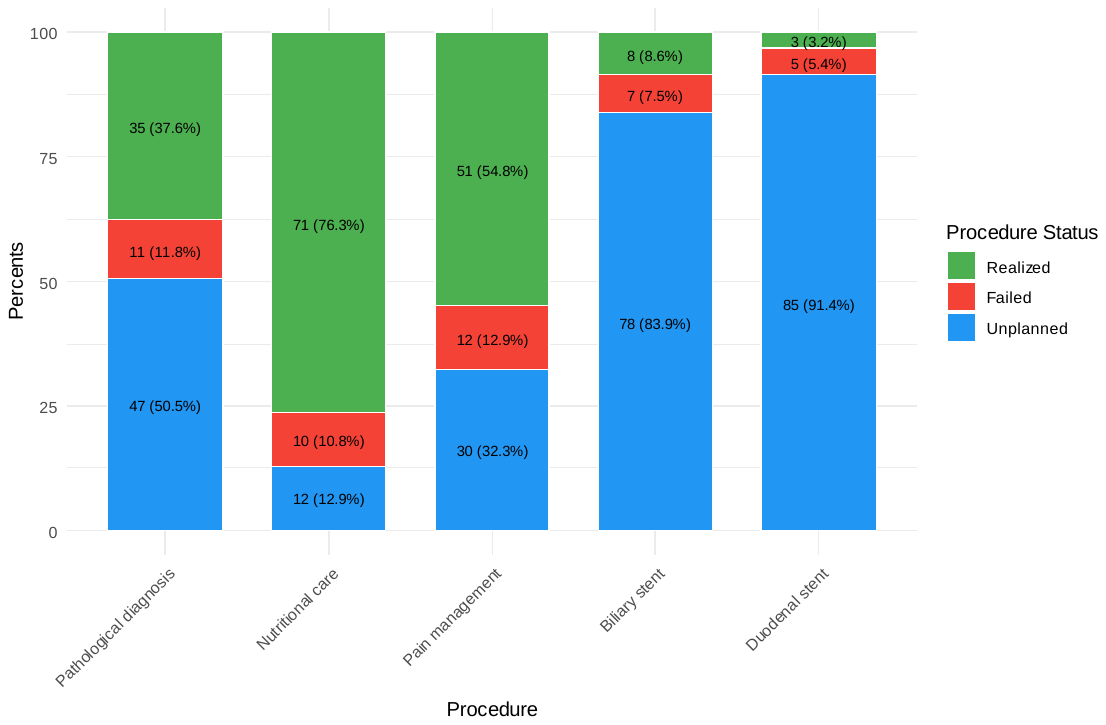
**A**

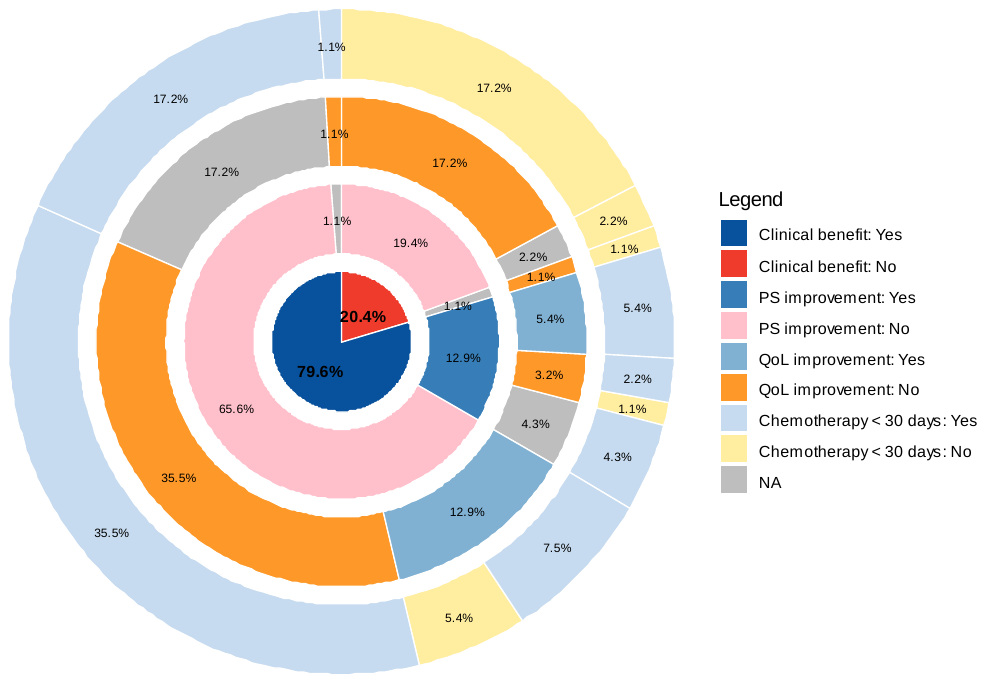

**B**

**Supplementary eFigure 3. Health-related quality of life (HRQoL) between baseline, V2 (second visit), and M1 (third visit at one month after visit 1 (V1).**

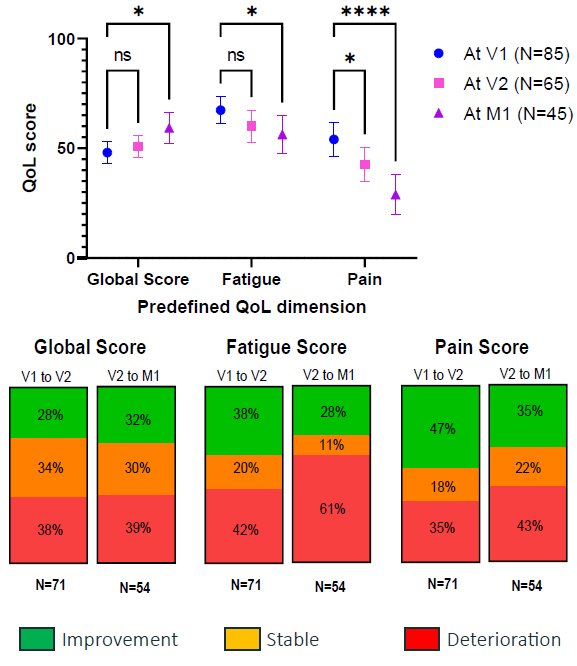

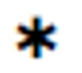

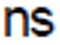

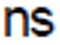

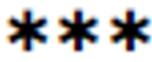

*: *P*<.05

**: *P*<.01

***: *P*<.001

**Supplementary Table 4.** Quality of life according to chemotherapy start in evaluable population for the primary endpoint analysis (n=99)

|  | **Global population (n=99)** | | | | | | | | | | | |
| --- | --- | --- | --- | --- | --- | --- | --- | --- | --- | --- | --- | --- |
|  | **V1** | | **V2** | | **M1** | | **V2-V1** | | **M1-V2** | | **M1-V1** | |
|  | n | Mean (SD) | n | Mean (SD) | n | Mean (SD) | n | Mean (IC95%) | n | Mean (IC95%) | n | Mean (IC95%) |
| **Global quality of life** | 85 | 48.0 (22.5) | 65 | 50.8 (19.9) | 45 | 59.3 (23.4) | 61 | 1.6 (-3.6, 6.9) | 41 | 5.3 (-1.8, 12.4) | 43 | 6.6 (-1.1, 14.3) |
| **Physical functioning** | 85 | 75.1 (25.3) | 66 | 72.5 (30.3) | 45 | 73.7 (26.9) | 62 | -5.6 (-12.9, 1.6) | 41 | -5.3 (-15.0, 4.4) | 43 | -8.9 (-18.2, 0.4) |
| **Emotional functioning** | 83 | 64.9 (28.3) | 65 | 71.0 (24.9) | 45 | 74.4 (27.7) | 59 | 5.4 (-1.0, 11.7) | 41 | 2.4 (-4.7, 9.5) | 41 | 11.4 (1.9, 20.8) |
| **Fatigue** | 85 | 67.4 (30.4) | 65 | 60.0 (29.9) | 45 | 56.3 (28.5) | 61 | -6.3 (-13.5, 0.9) | 41 | 1.6 (-10.4, 13.6) | 43 | -4.3 (-15.8, 7.3) |
| **Nausea & Vomiting** | 84 | 27.4 (34.0) | 66 | 27.3 (34.1) | 45 | 23.0 (34.7) | 61 | -3.3 (-13.3, 6.8) | 41 | 2.4 (-8.2, 13.1) | 42 | -5.6 (-17.7, 6.6) |
| **Pain** | 84 | 54.0 (35.1) | 65 | 42.6 (31.6) | 45 | 28.9 (30.6) | 61 | -10.4 (-17.8, -3.0) | 41 | -12.6 (-23.9, -1.3) | 43 | -19.4 (-30.8, -8.0) |
| **Dyspnea** | 85 | 23.5 (30.8) | 66 | 19.2 (26.2) | 45 | 22.2 (28.4) | 62 | 0.0 (-8.4, 8.4) | 41 | 13.0 (3.3, 22.7) | 43 | 6.2 (-3.1, 15.5) |
| **Insomnia** | 85 | 45.9 (38.5) | 66 | 32.8 (33.8) | 45 | 27.4 (32.0) | 62 | -10.8 (-20.3, -1.2) | 41 | -3.3 (-15.2, 8.7) | 43 | -14.0 (-27.6, -0.3) |
| **Appetite loss** | 85 | 71.4 (36.1) | 66 | 63.6 (34.5) | 45 | 55.5 (36.9) | 62 | -6.5 (-12.9, 0.0) | 41 | 1.6 (-11.5, 14.7) | 43 | -3.1 (-17.1, 10.8) |
| **Constipation** | 85 | 37.3 (38.3) | 64 | 38.0 (38.4) | 45 | 31.8 (33.3) | 60 | -1.7 (-13.2, 9.8) | 40 | -8.3 (-19.1, 2.4) | 43 | -7.0 (-19.6, 5.6) |
| **Chemotherapy start within 30 days (n=71)** | | | | | | | | | | | | |
| **Global quality of life** | 65 | 51.0 (22.8) | 50 | 51.3 (21.3) | 39 | 60.3 (23.4) | 47 | -0.4 (-6.1, 5.4) | 35 | 4.8 (-3.5, 13.0) | 37 | 6.8 (-1.3, 14.8) |
| **Physical functioning** | 65 | 76.4 (24.8) | 51 | 75.8 (30.1) | 39 | 75.2 (27.0) | 48 | -3.8 (-12.6, 5.0) | 35 | -5.2 (-15.2, 4.7) | 37 | -6.3 (-16.2, 3.6) |
| **Emotional functioning** | 63 | 64.0 (29.5) | 50 | 70.3 (25.3) | 39 | 73.9 (28.8) | 45 | 5.6 (-1.9, 13.0) | 35 | 3.3 (-4.6, 11.3) | 35 | 12.4 (2.5, 22.2) |
| **Fatigue** | 65 | 65.9 (30.8) | 50 | 59.0 (30.3) | 39 | 56.8 (27.5) | 47 | -6.0 (-13.8, 1.8) | 35 | 2.4 (-10.4, 15.1) | 37 | -3.2 (-15.4, 9.1) |
| **Nausea & Vomiting** | 64 | 28.1 (33.7) | 51 | 21.6 (30.4) | 39 | 17.9 (29.5) | 47 | -7.8 (-18.8, 3.2) | 35 | 0.9 (-11.3, 13.2) | 36 | -9.3 (-22.1, 3.6) |
| **Pain** | 64 | 52.9 (34.8) | 50 | 44.0 (31.5) | 39 | 28.2 (29.4) | 47 | -7.4 (-15.5, 0.6) | 35 | -13.8 (-25.8, -1.8) | 37 | -18.5 (-29.5, -7.4) |
| **Dyspnea** | 65 | 21.0 (29.8) | 51 | 17.0 (26.1) | 39 | 21.4 (27.0) | 48 | -0.7 (-9.5, 8.1) | 35 | 13.3 (4.5, 22.2) | 37 | 7.2 (-0.3, 14.7) |
| **Insomnia** | 65 | 46.7 (39.9) | 51 | 34.0 (33.7) | 39 | 27.3 (32.3) | 48 | -11.1 (-22.6, 0.4) | 35 | -2.9 (-13.8, 8.0) | 37 | -16.2 (-31.2, -1.3) |
| **Appetite loss** | 65 | 69.7 (37.6) | 51 | 60.8 (33.8) | 39 | 53.0 (35.6) | 48 | -8.3 (-15.4, -1.3) | 35 | 0.9 (-14.4, 16.3) | 37 | -4.5 (-20.3, 11.2) |
| **Constipation** | 65 | 36.9 (37.8) | 49 | 43.5 (38.6) | 39 | 33.3 (34.2) | 46 | 3.6 (-10.1, 17.3) | 34 | -9.8 (-21.5, 1.8) | 37 | -5.4 (-19.2, 8.3) |
| **No chemotherapy or chemotherapy start after 30 days (n=28)** | | | | | | | | | | | | |
| **Global quality of life** | 20 | 38.3 (18.8) | 15 | 48.9 (14.7) | 6 | 52.8 (24.5) | 14 | 8.3 (-5.1, 21.8) | 6 | 8.3 (-1.2, 17.9) | 6 | 5.6 (-28.9, 40.0) |
| **Physical functioning** | 20 | 70.8 (27.0) | 15 | 61.1 (29.3) | 6 | 63.9 (26.7) | 14 | -11.9 (-24.1, 0.3) | 6 | -5.6 (-50.7, 39.6) | 6 | -25.0 (-57.7, 7.7) |
| **Emotional functioning** | 20 | 67.5 (24.5) | 15 | 73.3 (24.2) | 6 | 77.8 (20.2) | 14 | 4.8 (-9.1, 18.6) | 6 | -2.8 (-23.2, 17.7) | 6 | 5.6 (-35.3, 46.5) |
| **Fatigue** | 20 | 72.5 (29.3) | 15 | 63.3 (29.0) | 6 | 52.8 (37.1) | 14 | -7.1 (-26.7, 12.4) | 6 | -2.8 (-51.5, 45.9) | 6 | -11.1 (-57.6, 35.4) |
| **Nausea & Vomiting** | 20 | 25.0 (35.7) | 15 | 46.7 (39.4) | 6 | 55.6 (50.2) | 14 | 11.9 (-12.7, 36.5) | 6 | 11.1 (-6.9, 29.1) | 6 | 16.7 (-26.2, 59.5) |
| **Pain** | 20 | 57.5 (36.9) | 15 | 37.8 (32.4) | 6 | 33.3 (40.8) | 14 | -20.2 (-39.2, -1.3) | 6 | -5.5 (-50.7, 39.6) | 6 | -25.0 (-88.3, 38.3) |
| **Dyspnea** | 20 | 31.7 (33.3) | 15 | 26.7 (25.8) | 6 | 27.8 (39.0) | 14 | 2.4 (-22.0, 26.8) | 6 | 11.1 (-46.0, 68.2) | 6 | 0.0 (-66.4, 66.4) |
| **Insomnia** | 20 | 43.3 (34.4) | 15 | 28.9 (35.3) | 6 | 27.8 (32.8) | 14 | -9.5 (-27.1, 8.1) | 6 | -5.6 (-77.0, 65.9) | 6 | 0.0 (-44.3, 44.3) |
| **Appetite loss** | 20 | 76.7 (30.8) | 15 | 73.3 (36.1) | 6 | 72.2 (44.3) | 14 | 0.0 (-16.9, 16.9) | 6 | 5.6 (-8.7, 19.8) | 6 | 5.6 (-28.8, 40.0) |
| **Constipation** | 20 | 38.3 (40.9) | 15 | 20.0 (32.9) | 6 | 22.2 (27.2) | 14 | -19.1 (-38.6, 0.5) | 6 | 0.0 (-38.3, 38.3) | 6 | -16.7 (-59.5, 26.2) |

For global quality of life, physical functioning and emotional functioning, a higher score represents a higher ("better") level of functioning. For symptoms, a higher score represents a higher ("worse") level of symptoms.
